# Modeling epidemiological patterns of smallpox in Copenhagen in the 19*^th^* century after the introduction of the vaccine

**DOI:** 10.64898/2026.01.05.26343436

**Authors:** Andreas Eilersen, Søren K. Poder, Bryan T. Grenfell, Kim Sneppen, Lone Simonsen

## Abstract

In 1798, Jenner’s smallpox vaccine made it possible to prevent the deadliest of childhood diseases. In Denmark the vaccine was used from 1801, and by 1810 a mandatory 1-dose childhood vaccination program was instituted, free of charge. As proof of vaccination (or natural immunity) was required for church confirmation, most children were vaccinated and smallpox disappeared from Copenhagen after 1810 [1]. After a 14-year “honeymoon period”, smallpox returned in 1824 with a new face: a milder disease in mostly young adults. Here we investigate the effects of smallpox vaccination on the epidemic patterns throughout the post-honeymoon era (1824-1875). We accessed data from the hospital “Søkvæsthuset” where all smallpox cases, mild and severe, were hospitalized during 1824-1835 in order to contain the outbreak. We identified ∼ 3000 smallpox cases in four separate epidemics occurring during this period and accessed data on the age and vaccination status of patients where available. In addition, some information on the number and age distribution of severe smallpox cases and on mortality for outbreaks in the 1860’s and 70’s is available. We used a mechanistic model (SEIR) to assess factors explaining the return of smallpox, and the changing mean age of cases. We considered vaccination coverage and effectiveness, duration of vaccination-induced immunity, and the fate of the “lost generation” of persons born around 1800, too early to get vaccinated and too late to have been infected with smallpox. Our model tracks well the disappearance and return of smallpox in 1824, its age patterns, and the interval between epidemic peaks. Our study shows that vaccine waning after 25 years on average was likely the primary reason explaining the return of smallpox and the change in its epidemiology in the vaccine era.

## Introduction

Smallpox was a devastating disease that profoundly affected past populations and societies due to its high mortality and the fact that nearly everyone was infected in childhood [2, 3]. As the first disease to be controlled by a vaccine – and by 1979, to be eradicated globally – it also represents an iconic public health success that shaped modern medicine and inspired the global Expanded Programme on Immunization (EPI).

Smallpox was caused by Variola, a virus which was transmitted through the respiratory route, though infection could also occur via direct contact with infected bodily fluids, contaminated bedding or clothing. After infection, an incubation period of typically 10–14 days passed before general symptoms such as fever appeared [4]. This prodromal phase was followed by the onset of a macular rash that developed into pustules within four to seven days of the appearance of the macules. The pustules then required five to eight days to rupture and form scabs. The infectious period generally ended once the scabs began to fall off, “usually by the second week of the eruption” [2].

Because smallpox was such a deadly disease, methods to control it were pursued even in pre-industrial times. It was discovered that immunity could be induced by deliberately exposing children to small amounts of infectious material through the skin, ideally resulting in a mild infection. This method, known as variolation, spread to Europe in the 18*^th^* century [5] from Asia where the practice dated back to at least 1500 AD [6]. In 1798, Edward Jenner developed a safer vaccine based on similar principles but using the related, much less virulent cowpox virus to induce immunity [7].

The introduction of the smallpox vaccine was met with considerable enthusiasm, particularly within the medical community and among the socio-economic elite [1]. Across multiple countries, including Denmark, vaccination initiatives were rapidly implemented. The public health infrastructure was still in its early stages. Nevertheless, due to a past variolation program foundational infrastructure for vaccine administration was in place by late 1801 when the smallpox vaccine was introduced in Denmark [8, 9]. In the first decade, smallpox vaccination was voluntary but by 1810, the vaccine was made semi-mandatory and was provided free of charge. The enforcement involved requiring vaccination as a condition for church confirmation, which was socially obligatory. As a result vaccination coverage after 1810 was high [1, 10].

Following the success of the vaccination program, smallpox incidence was greatly reduced, and the disease was effectively eliminated from Copenhagen after 1810. It remained absent from the city during a 14-year “honeymoon period” before returning in 1824 with a different clinical presentation and epidemiology. Now the disease was milder and predominantly affected young adults [1, 11]. Older individuals, particularly those born before 1795, had natural immunity and were rarely among the medically attended cases [11–13].

Denmark, and especially Copenhagen, is an ideal setting for further research into the epidemiological patterns of smallpox in the early vaccine era due to the existence of extensive records of both fatal and non-fatal smallpox cases compiled by physicians. In these records, the vaccination uptake and circumstances were carefully described [1, 10, 14]. By integrating these historical data with mathematical modeling, we address several open questions, such as identifying the main factors explaining the observed post-vaccine age patterns of smallpox.

Firstly, we aim to identify factors that contributed to the resurgence of smallpox. As natural immunity became less prevalent due to a reduction in smallpox incidence, the population increasingly relied on vaccine-induced immunity. We will investigate four potential factors that enabled smallpox to bypass this immunity: (1) waning vaccine effectiveness over time; (2) the presence of a “lost generation” born too late to have acquired natural immunity and too early have been vaccinated in childhood; (3) vaccine failure, occurring when the vaccine did not induce immunity against smallpox; and (4) vaccine leakiness, i.e., the vaccine reduced but did not eliminate the risk of getting infected regardless of time since vaccination (see e.g. [15]). These factors are not mutually incompatible and may all have contributed to the observed epidemic resurgence and changing manifestation of smallpox.

Therefore, we developed a dynamic transmission model calibrated to the situation in the pre-vaccination era and run to an endemic equilibrium with smallpox as a childhood disease. Using the resulting immunity distribution as our starting condition, we then simulated the vaccination rollout, honeymoon period and post-honeymoon epidemic era. We compared the resulting theoretical predictions with empirical observations made during the 1824, 1828-30, and 1835 smallpox outbreaks in Copenhagen.

## Materials and methods

### Data on vaccine uptake

Data on vaccine coverage were derived from official records maintained by public health authorities [16] and from counts by contemporary doctors [11–13]. Each child received a single dose of the vaccine, with revaccination not becoming common until the 1870’s [13, 17]. Strict enforcement of the vaccination mandate, coupled with systematic record-keeping of vaccination certificates and registries of immunized individuals provided reliable data on coverage. Vaccine uptake increased over time during the rollout in 1802-1811, following an uneven pattern that reflected higher vaccination rates during epidemic years. Using these data in conjunction with birth records, we estimate that approximately 50 % of children born in Copenhagen after 1800 were vaccinated by 1810. Following the introduction of the mandatory, free vaccination program in 1810, strong social pressure contributed to widespread uptake, and from then on around 80 % of each birth cohort were vaccinated. Assuming 90 % of children survived long enough for vaccination to be a relevant possibility [18], we estimate an effective coverage of approximately 90 % of surviving children from 1811. The vaccine coverage over time is estimated by dividing the number of doses administered by the size of the birth cohort of the previous year. A timeseries of these data is found in the supplement (fig. S2(b)). Vaccination coverage remained high throughout the study period, as anti-vaccination movements in Denmark did not gain significant traction until after 1900 [9]. Revaccination was debated over the years, but it was not free of charge and few were revaccinated, mostly army recruits [13]. In 1871, an even more stringent vaccination law was enacted, requiring verification of vaccination status for school entry and strongly encouraging re-vaccination during outbreaks [17]. These enhanced practices likely contributed to the eventual elimination of smallpox in Denmark by the close of the 19*^th^* century.

### Smallpox case records

Data on the historical smallpox epidemics discussed here were obtained from archival records of smallpox hospitalizations in 1824-35, collected by the smallpox quarantine hospital Søkvæsthuset and accessed via the Danish National Archives [19, 20], medical reports (Medicinalberetningerne) [21]. Data on cases by vaccination status were found in contemporary works by doctors Møhl, Hoppe, and Wendt [11–13]. [11] and [13] in particular contain case counts for 1824 and 1835 stratified by vaccination status, and therefore we use these as the main basis for the age distributions shown in fig. 2. The data given by Hoppe [12] contain counts of vaccinated, but not unvaccinated cases. We calculate the number of unvaccinated cases in 1828-30 by subtracting the vaccinated case counts of Hoppe from the total case counts from Søkvæsthuset. It should also be noted that there is a small discrepancy between the case counts reported by Hoppe [12] and the Søkvæsthuset data [20], since for some age groups, Hoppe reports up to six more cases in the vaccinated alone than the total number of cases in the same age group as reported by Søkvæsthuset. Nonetheless, we still use these data, as the discrepancy is quite small and likely due to differences in registration. We concentrate on the age distributions from 1824, 1828-30, and 1835, since for other outbreaks we only have the ages of patients, but not the vaccination status.

**Figure 1:**
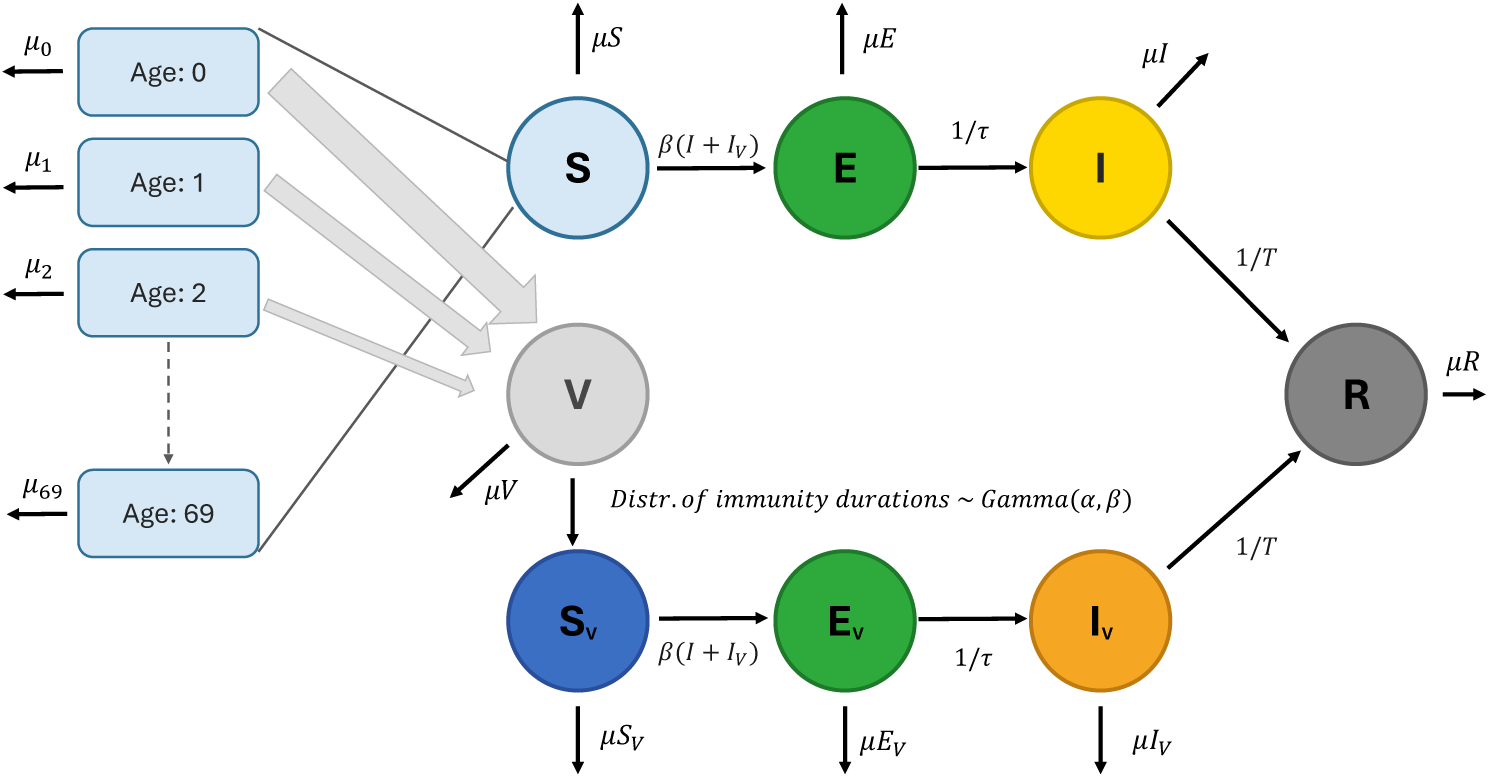
A diagram of the model used to simulate smallpox dynamics in the vaccine era. Vaccine immunity waning times are assumed to be Gamma distributed with a form factor of *α* = 4 and a mean of *τ_V_*. The vaccine provides 100 % protection before waning. Births and immigration are not shown in the diagram. For each year of life up to age *m*, a certain fraction of the remaining susceptible children are vaccinated as indicated by the gradually narrower arrows from the youngest age groups to the *V* compartment. At age *m*, the vaccinated fraction is *v_obs_*(*t*), which reaches *v_max_* = 90% by 1811. The maximum age at vaccination, *m*, is assumed to decline linearly until reaching one year at the outbreak in 1824, based on data from [16], visualised in supplementary fig. S2(a).

**Figure 2:**
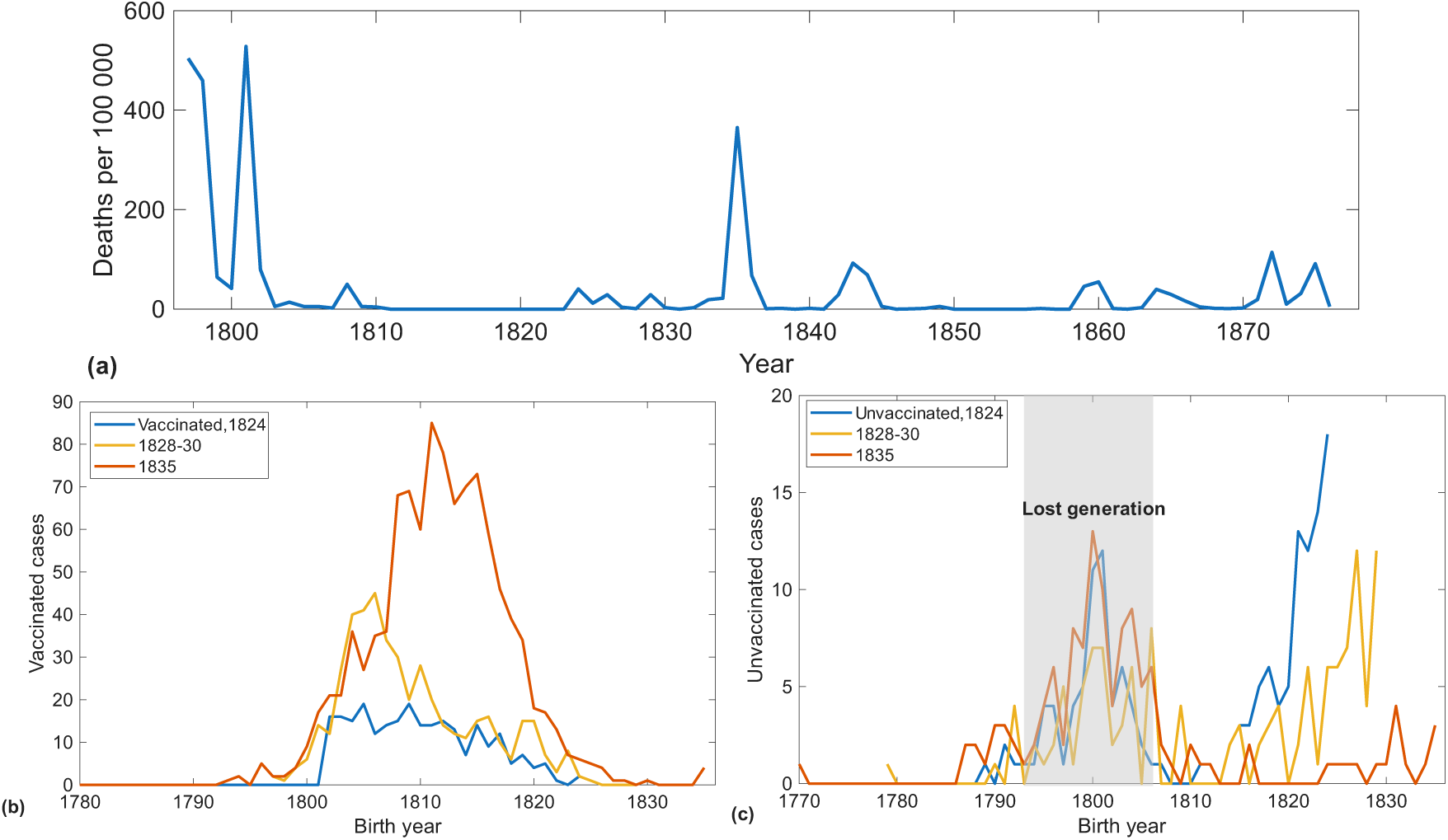
Observational data from the vaccination campaign and smallpox outbreaks in the 1800’s. (a) Time series of smallpox deaths per 100 000 inhabitants in Copenhagen from 1796 to 1877. (b) Number of vaccinated smallpox cases in the 1824 and 1835 outbreaks and in the years 1828-30 by birth year. (c) The same for vaccinated patients. Data from [11–13, 21, 23].

The data from Søkvæsthuset [20] cover four outbreaks that occurred between 1824 and 1835 when smallpox re-emerged in smaller, milder epidemics, predominantly affecting adults. During 1824-1835, the ∼ 3000 records represent all medically attended cases in the city, regardless of severity. This comprehensive data collection was possible due to quarantine measures requiring hospitalization of all infected at Søkvæsthuset [1, 11, 12]. However, after May 1835, these quarantine policies were discontinued as smallpox was no longer regarded as a major public health threat [13]. As a result, any records from May 1835 onwards represent a subset consisting of severe smallpox cases, as patients could now be treated at any hospital in Copenhagen, and as many mild cases were no longer documented. For the outbreaks in Copenhagen after 1835, we therefore relied mainly on mortality time series, which we have assembled from the whole 19*^th^* century from [21–23]; these are presented in figs. 2(a) and 4(b), where the time series is compared with simulation results. An overview of the age distribution and vaccination status of those hospitalized in the 1871-72 outbreak can be found in the supplement (fig. S12). These data are not part of the main analysis due to the large discrepancy in hospitalization policies which means that the later data no longer include any mild cases.

### The model

Based on the clinical description from [2], we constructed a dynamical model of the disease using ordinary differential equations (ODE’s). These each describe the rate of change of a subpopulation, e.g., the susceptible or infected. Since smallpox clearly has a separate latent and infectious period, we use the SEIR (susceptible-exposed-infected-recovered) framework. Thus, we divide the population into four compartments based on their susceptibility to the disease and infectivity. Only those in the infectious (*I*) compartment can pass on the disease. Similar models have been proposed for smallpox before, mostly over the last decades in the context of the bioterrorist risk posed by the disease [3, 24–26]. Since age played a key role in smallpox dynamics, we divide the model population into 70 age groups of a one-year width each, assuming that populations older than 70 are small enough to play a negligible role in transmission. At discrete one-year intervals, we move up the entire population to the next age group. Each age group has an age-specific background mortality rate, and deaths are countered by an equal number of births, keeping the population constant. Modeled individuals are assumed to die when aging out of the oldest age group.

We run a simulation in the absence of vaccination for 70 years to obtain an equilibrium distribution of natural immunity where every age group has been exposed to smallpox. This immunity distribution is then used as the initial immunity distribution *R_i_*(0) of every post-vaccine simulation, saving time by allowing us to only simulate the post-vaccine era when testing the effect of different vaccination parameters. We start the post-vaccine simulation with a single infected person in the youngest age group, and initiate the vaccination rollout in the immediate aftermath of an outbreak as was historically the case [1]. To achieve this, the main simulation starts five years before the first vaccinations, corresponding roughly to the year 1796. See supplementary fig. S4 for a full simulated timeseries from 1726 to 1876 compared with a smallpox mortality timeseries.

With smallpox established in the population, we vaccinate the simulated population in childhood with an increasing coverage that reaches a steady level of 90 % from 1811. We assume that vaccine immunity initially gives 100 % protection from infection. Immunity after infection is assumed to be lifelong but vaccine-induced immunity wanes after some time. The duration of immunity in each individual we assume to be Gamma distributed. We expand upon the reasoning behind this distribution in the supplement. We vary the mean duration of vaccine immunity and assess which value results in the best fit with data.

To investigate the effect of an imperfect vaccine on the population, we introduce four other compartments: a vaccinated immune compartment, a vaccinated susceptible compartment for those with waning vaccine protection against infection, and vaccinated exposed and infectious compartments for breakthrough infections. To achieve a Gamma distributed vaccine immunity duration, the vaccinated immune compartment *V* is further divided into four sub-compartments *V*_0_*…V*_3_, through which the vaccinated pass with exponential rates of 4*/τ_V_*, giving a mean duration of vaccine immunity of *τ_V_*. Some percentage of children are assumed to be vaccinated each year until they reach an age of *m* years. This age declines until 1824, when we have data indicating that a majority of children were vaccinated before their first birthday. The model equations thus become

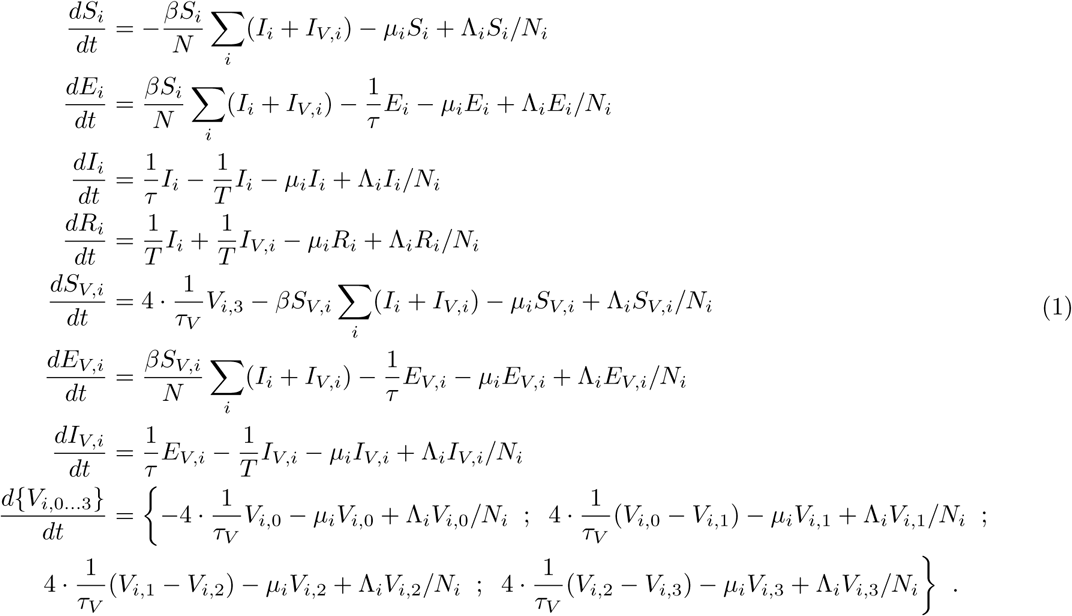

Here, *S_i_* are susceptibles of age class *i*, *E_i_* are exposed and non-infectious, *I_i_* are infectious, *R_i_* recovered and immune, *V_i,j_* vaccinated and immune, *S_V,i_* vaccinated and susceptible, and *E_V,i_*, *I_V,i_* are exposed and infectious vaccinated, i.e., breakthrough infections. Vaccinated people in the *V_i,j_*compartments are assumed to have full sterilizing immunity, while vaccinated susceptible people (*S_V_*) are as vulnerable as the unvaccinated. Some fraction of the births are vaccinated and thus moved into the *V*_1,0_, and initially the same is true for the youngest age groups up to age *m*. Maternal immunity in the first months of life may be present in smallpox but is ignored in the model [27]. The processes of birth and vaccination are implemented numerically and are thus not shown in the presented equations.

*β* is the transmission rate, *τ* is the duration of the latent period, and *T* is the duration of the infectious period, here both taken to be 12 days. This is relatively short given the range for both the latent and infectious periods, but given the observed variability of the periods, rounding does not cause any significant problems not already present due to the uncertainty of the empirical mean duration. We set the daily transmission rate to *β* = 0.58, giving an *R*_0_ of 7. This is at the upper end of the estimates given by [28, 29]. The high value is chosen as Copenhagen at the time was very densely populated and a higher *R*_0_-value results in a more realistic age distribution of smallpox patients, specifically a sufficiently high fraction of cases occurring among children. In the supplement (fig. S11) we test the sensitivity of the model to this assumption. The vaccine immunity duration, *τ_V_*, is unknown and in this paper we attempt to determine it by fitting the model to data. An overview of all parameters used can be found in Table 1.

**Table 1:** Parameters used in the presented model.

| Parameter | Definition | Observed range | Values used | Sources |
| --- | --- | --- | --- | --- |
| $R_0$ | Basic reproduction number | 5-7 | 7 | [28, 29, 33] |
| $\tau$ | Latent period | 9-23 days | 12 days | [2, 4] |
| $T$ | Infectious period | 12-20 days | 12 days | [2, 4] |
| $\beta$ | Transmission rate | - | 0.5833 /infectious/day | $R_0/T$ |
| $m(t)$ | Max. age at vaccination | 1-14 years, decreasing | $m(t_{start} < t < 1824) = 8 - \frac{(t-t_{start}) \cdot (8-1)}{1824-t_{start}}$<br>$m(t \geq 1824) = 1$ | [16] |
| $v_{obs}(t \geq 1811)$ | Observed vac. coverage after vaccine mandate | $\approx 90$ % | 90 % | [13, 16] |
| $v_{obs}(t < 1811)$ | Observed vac. coverage before mandate | Mean $\approx 50$ % | Vac. protocol data (see supplement) | [13, 16] |
| $\tau_V$ | Mean duration of vaccine-induced immunity | 3-60 years (3-20 for transmission-blocking) | Fitted parameter | [34–37] |
| $\text{Gamma}(\alpha, \beta)$ | Distribution of vaccine waning times | - | $\alpha = 4$<br>$\beta = 4/\tau_V$ | Assumed |
| $\Lambda_i$ and $\mu_i$ | Migration and death rates | Infant mortality 15 % for *all of Denmark* | See supplement (one rate per cohort) | [30, 31] |
| $N$ and $N_i$ | Total population /pop. of age class $i$ | Total: 121 868 | $N = 121868$,<br>$N_i$: See fig. S3 | [30] |
| Reintroduction interval | - | Unknown | 14 days | Assumed (see [1]) |

*µ_i_* is an age-dependent death rate, which however is independent of disease, as making demographic processes dependent on epidemic dynamics would greatly complicate the model. We see this omission as justified, as our data indicate a relatively low smallpox mortality relative to the population size in the vaccination era. Every death, including those modeled by having the oldest age group “age out” of the system is balanced by a birth into the compartment *S*_1_. Migration is modeled by adding a number Λ*_i_* to each of the age groups between 15 and 25 years of age. We add immigration in these age groups to emulate the demography of Copenhagen at the time, in which the 15-25-year-olds are strongly overrepresented, likely due to immigration. The population pyramid from the 1834 census [30] and that used in our model can be found in supplementary fig. S3. The immigrants are distributed evenly over all compartments *S, E, I,…*, hence why we multiply by the factor [*compartment population*]*/N_i_*, where *N_i_* is the total population of the age group. Immigration is balanced by an equal reduction in births.

Death and migration rates *µ_i_*and Λ*_i_* are estimated by assuming an urban infant mortality that lets the size of the youngest cohort match the demographic data. For this, an infant mortality of 180/1000 is needed, which is high, but we would argue that it is realistic. Sources estimate a lower infant mortality of 150/1000, [31], but this includes rural areas, and urban populations are likely to have been subject to the “urban mortality penalty” [32]. We would also argue that it is not inconsistent with our assumption that 10 % of children die before vaccination becomes a possibility, as the 18 % includes everyone who dies before their first birthday, whereas the 10 % only includes those who die before vaccination, and data show that many vaccinations occurred in those less than one year old. With this rate fixed, we used the Claude Sonnet 4.6 LLM to estimate a set of birth and death rates resulting in a steady-state demography matching the one observed in 1834 [30]. By running the simulation with the rates “guessed” in this way, we verified that they produced a demographic population profile that matched the one observed in the 1834 census (see supplement, fig. S3). The exact values of *µ_i_* and Λ*_i_* used can be seen in the supplement.

The initial conditions are *S_i_*(0) = *N_i_*(0) − *R_i_*(0) − 1 and *I*_1_(0) = 1. The total population of each age cohort, *N_i_*(0), is taken from the 1834 census (see fig. S3). From this, we get a total population of 121 868 individuals. All populations except *S, I, R* start at zero.

The vaccination program is modeled as follows. To emulate the initial rollout, we use the observed vaccination rates *v_obs_*(*t*) for Copenhagen during the first 10 years after the introduction of the vaccine, before it was made mandatory. These, we take from Wendt [13]. After the introduction of the mandate, *v_obs_* is assumed constant at a standard value of *v_max_*= 90%. For the first *m* years of life, a fraction *f_Y_* = 1 − (1 − *v_obs_*)^1*/m*^ of all susceptible children are vaccinated at each “birthday”. This ensures that the vaccinated fraction in the absence of natural immunity reaches *v_obs_* at age *m*. The maximum age at vaccination, *m*, starts at 8 years and is lowered linearly until reaching *m* = 1 in year 28 of the simulation (corresponding roughly to the calendar year 1824), after which the data show that the vast majority of children were vaccinated in their first year of life (see [16] and supplementary fig. S2(a)). When modeling the leaky vaccine, the *V*_0,1,2,3_ and *S_V_* compartments are merged into a single vaccinated compartment. Vaccinated individuals are susceptible to infection, but with a *β* = *β_V_*which is lower than for the unvaccinated. Infected vaccinated are just as infectious as infected unvaccinated. For the full details of this model, see the uploaded code (DOI: 10.6084/m9.figshare.28182581).

Finally, as the possibility of stochastic disease extinction has a significant effect on real epidemics [38], we also incorporate it in this model by giving the epidemic a probability of 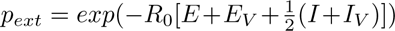 of going extinct for each infectious period *T*. This is equivalent to the Poisson probability of the available exposed and infectious individuals not producing any new infections before recovering. The factor of ^1^ is introduced since a randomly chosen infected individual will on average be halfway through their infectious period at a given random time point. A single new case is introduced randomly with immigration into age group 15 at a rate of 24 cases/year, equivalent to one reintroduction per two weeks. We unfortunately have no sources available that provide a solid realistic estimate for this parameter, but as Copenhagen at the time was a well-connected port city, we expect reintroductions to have been relatively frequent. Furthermore, while the reintroduction frequency does change the frequency of epidemics, the latter depends much more strongly on the amount of susceptibles present. Reintroducing the disease into the same age group every time may skew the age distribution if numbers of cases are very small, but during an outbreak it causes a negligible error.

## Results

### Observational data, 1824-35

Initially, we accessed the records of smallpox cases made during the outbreak period 1824-35. We show the most important data in fig. 2 [11–13, 20]. Further details on vaccination coverage and age at vaccination can be found in the supplementary fig. S2 [16]. Some notable facts can already be gleaned from these observations. The time series of smallpox mortality per 100 000 in panel (a) clearly shows the honeymoon period and the small outbreaks between 1824 and 1835 that we study. In 2(b,c) we plot the number of unvaccinated and vaccinated cases in 1824, 1828-30, and 1835, by birth year. For the unvaccinated we have shaded the period from the early 1790’s to just before 1810 to indicate the “lost generation”, i.e., the birth cohorts born too early to be vaccinated but too late to be infected in childhood. As hypothesized, persons born in this decade were strongly overrepresented among the unvaccinated smallpox cases in both the 1824, 1828-30, and 1835 outbreaks. In 1824, and to a lesser extent 1828-30, the youngest age cohorts born after 1815 are also overrepresented among the unvaccinated cases. This is not the case in 1835, consistent with vaccinations gradually occurring at an earlier and earlier age, as contemporary vaccination protocols indicate (see supplementary fig. S2(a) and [16]).

Furthermore, as seen in fig. 2(b), the distribution of vaccinated cases by birth year indicates some vaccine immunity waning. In the 1824 outbreak, we see that the number of cases in the vaccinated is highest in the earliest vaccinated cohorts (born around 1801). The same is the case in the 1828-30 data, but here the peak in the oldest cohorts is higher. In 1835, the peak has shifted towards those born around 1810. The vast majority of children born in 1810 or later would have been vaccinated in early childhood and the cases in 1835 were thus vaccinated 20-25 years previously. Note that due to the change in quarantine policy, the 1835 data only cover the months of May-December, and only one of the multiple hospitals which were by then open to smallpox patients [13].

From these three data sets we asses that the timescale of the waning is around 25 years. The main argument for this is the fact that the vaccinated case counts by birth year (fig. 2(b)) and by age (fig. S8(a)) spike in the cohorts who were of vaccination age 25 years before the outbreak in question. Relatedly, we observe very few cases in the youngest, recently vaccinated age groups, indicating a relatively consistent vaccine immunity duration and speaking against the hypotheses of a leaky vaccine or vaccine failure.

### Analysis

Having studied these observational data for three epidemics during the early post-honeymoon era, we compare them to the simulated dynamics in fig. 3. Fig. 3(a,b) shows the simulated age distributions of cases in the unvaccinated and vaccinated over time for a vaccine immunity duration of 25 years and 90 % coverage. From this, we see that outbreaks are predicted to occur about once every six years on average after a honeymoon period of 15 years duration. This is very similar to the observed honeymoon period of 14 years and the observed outbreak interval of 5.67 years (depending on counting criteria) in the post-honeymoon period. The most affected age groups also appear to gradually get older, as observed in reality. Early on, cases in the unvaccinated population dominate (panel a, red), but later in the simulation, we predict breakthrough cases in the vaccinated (panel b, blue) to displace them. Late in the simulated period, there are few remainining unvaccinated cases, focused in the youngest age groups. It is notable that in the data of fig. 2(b,c), cases in the vaccinated dominate already in the earliest post-honeymoon outbreaks, but this does not happen to the same extent in the simulations of 3(a,b).

**Figure 3:**
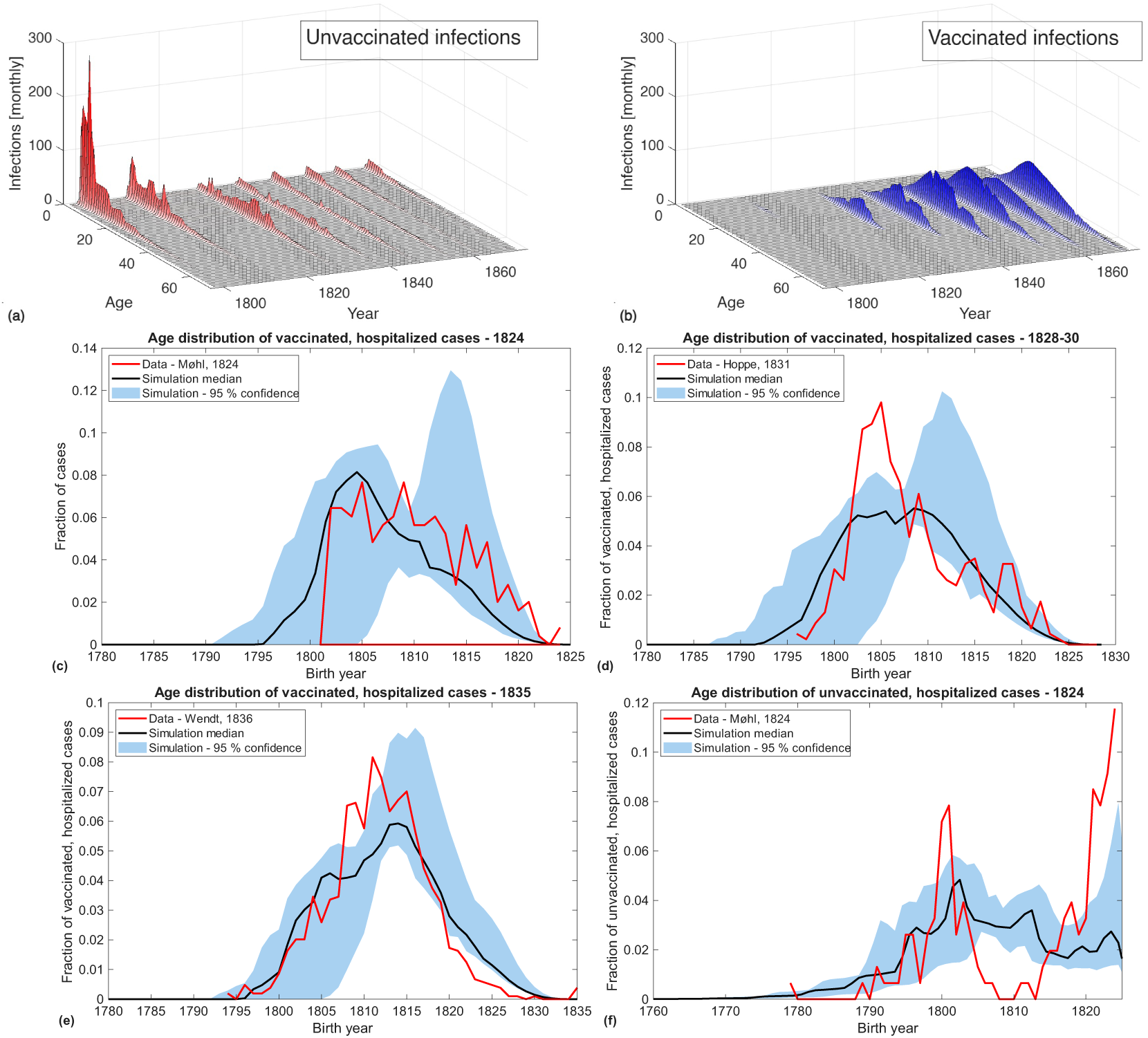
Simulations of epidemic dynamics and birth year distributions after vaccine rollout. (a,b) Monthly infections by age over time in the unvaccinated (red) and vaccinated (blue) population. (b) Same for the vaccinated population. (c-f) The fraction of cases in the vaccinated who were born in a given year, compared with the simulated fraction of infections who were born in the same year. (c) shows this distribution for vaccinated cases/infections in the 1824 outbreak, (d) for the vaccinated cases in 1828-30, (e) for the vaccinated cases in 1835, and (f) for the *unvaccinated* cases in 1824. Comparisons between simulations and data for the unvaccinated populations are found in supplementary fig. S7. All simulations were done using a vaccine immunity duration of 25 years and a coverage of 90 %.

Fig. 3(c-f) compares simulated age distributions of infections with those of cases in the 1824 outbreak, in the years 1828-30, and in the 1835 outbreak. Panels (c-e) show data for vaccinated cases, while (f) shows unvaccinated cases in 1824. These are compared with the simulated age distributions of infections in the first and second post-honeymoon outbreak for a vaccine coverage of 90 % and a vaccine immunity duration of 25 years. We deal with the stochastic variability of the model by running the simulation 20 times, allowing us to obtain a 95 % confidence interval of the measured quantities. The plots show the fraction of cases (or infections, in the case of the simulations) of a given vaccination status that were born in a given year. They are thus normalized such that they all sum up to 1.

In terms of timing, the simulated outbreaks roughly correspond to the historical 1824 and 1835 outbreaks. However, in the simulation there is no outbreak corresponding to that in 1828-30. We handle this by comparing the data from those years to the simulated age distribution of infections from a four-year period between the two first post-honeymoon outbreaks. The four-year period is chosen instead of the two-year period of the data, as a shorter period between outbreaks would contain very few cases, letting the model’s periodic reintroduction of a single smallpox infection in the migrating young adult population skew the age statistics. For all three vaccinated birth year distributions (c-e), representing the outbreaks in 1824, 1828, and 1835, the simulations fit the birth year patterns well. For unvaccinated cases (panel (f)), the simulated birth year distribution deviates further from the observed one, but nonetheless has a noticeable peak around 1800, meaning that it captures the effect of the lost generation. The coefficients of determination measuring how well the simulation fits all available age distribution data can be found in the supplementary fig. S5. Note however that age distributions and birth-year distributions do not correspond one-to-one, as the exact timing of the epidemics will affect the age of the infected in each outbreak.

We likewise compare the change in mean age of observed cases in the post-honeymoon period with the simulated mean age of infected. In fig. 4(a) we see a graph of the predicted mean ages of infected given a vaccine coverage of 90 % and a vaccine immunity duration of 25 years. The mean ages observed in data are shown as red dots. The *R*^2^-value of the fit is 0.76. In panel (b) we show a comparison between the observed time series of smallpox deaths (left y-axis) and the simulated time series of infections (right y-axis), both measured per year per 100 000 inhabitants. The simulated time series is a single run of a simulation with stochastic extinction and therefore the timing of the peaks is likely to vary between runs. Although we have not attempted to get the exact timing of epidemic peaks to match observations, the simulated outbreaks match the data quite well.

**Figure 4:**
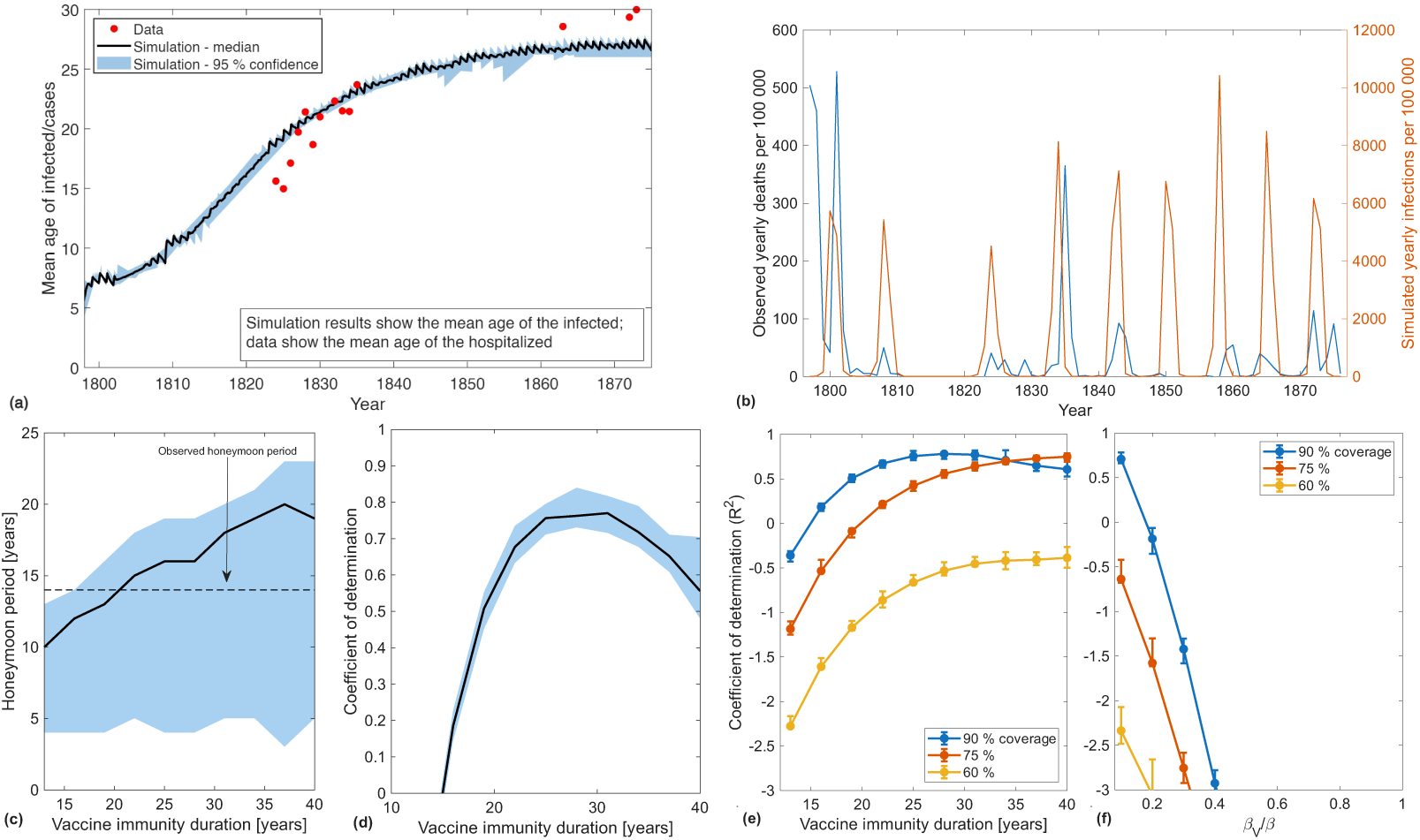
Observed and simulated timeseries of number of cases and goodness-of-fit values. (a) A comparison of the simulated mean age as a function of time (black curve) with the observed mean ages (red dots). We have removed the years 1831 and 1864, as they were outliers with few cases. (b) A comparison between the real time series of the smallpox mortality and the simulated time series of infection incidence. (c) shows the model’s prediction of the duration of the honeymoon period and (d) the values of the coefficient of determination (*R*^2^) for fits of the mean age of infected over time to data when varying vaccine immunity duration. An *R*^2^ close to 1 indicates a good fit. In (e,f) we compare the *R*^2^ values of the model with waning vaccine immunity with those of a model with a leaky vaccine, showing that the best fits (*R*^2^ closest to 1) is produced by the waning vaccine model for a vaccine immunity duration of 25-30 years. The *R*^2^ curve of panel (d) and the blue curve of panel (e) are not exactly identical due to stochastic variation. Shaded blue areas and error bars both show 95 % confidence intervals. Where not stated otherwise, vaccine immunity duration is 25 years and coverage is 90 %.

Fig. 4(c-d) show the honeymoon period duration predicted by the simulation and the coefficient of determination of the mean age time series fitted to data. These results are shown for varying vaccine immunity duration and a coverage of 90 %. The black lines show the median of the simulated results, while the shaded blue areas show the 95 % confidence intervals. We observe that the most accurate honeymoon period duration is obtained when setting the vaccine duration to 20 years, while the best fit of the mean age data (i.e., that with an *R*^2^ closest to 1) is achieved in the 20-30 year range.

To evaluate the hypothesis that many vaccinations failed to induce immunity we vary the vaccine coverage used in the model from the observed 90 % down to 60 %. We do this since if some vaccinations were non-immunizing this would from a disease-dynamical perspective be equivalent to a lower vaccine coverage. The results of this is shown in fig. 4(e,f). Here we compare the goodness-of-fit measured by *R*^2^ between the mean age data and the simulation results for the waning vaccine and leaky vaccine models. For the leaky vaccine model (panel (f)) we vary the transmission rate of the vaccinated *β_V_* and the vaccination coverage, while for the waning vaccine model (panel (e)) we vary the waning time as in panels (c,d). We see that the best fits for the mean age of infected as a function of time are produced by the waning-vaccine model when vaccine coverage is close to 90 % and immunity duration is between 20 and 30 years. Measured by the *R*^2^ value of the fit, the leaky vaccine model might explain the mean age data almost as well as the waning vaccine model if the vaccine reduces infectivity by 90 % (*β_V_ /β* = 0.1), but not otherwise. However, we rule it out based on the observation that very few vaccinated children contracted smallpox (fig. 2(b) and S8(a)). If a vaccine was uniformly leaky at every time after administration, we would expect to see some cases in the most recently vaccinated children, but these are notably absent.

In the supplement (fig. S5), we include plots of *R*^2^ for fits between the simulated age distributions in 1824 and 1835 for the waning and leaky vaccine models and varying parameters. The fits of vaccinated age distributions broadly corroborate our findings above. However, the unvaccinated age distributions generally have a relatively poor fit between data and simulation, as is also illustrated by fig. 3(f). We discuss possible explanations for this below.

From the comparison between observational data and simulations shown in fig. 4, we can now tentatively conclude which of the investigated factors played the biggest role in the return of smallpox and subsequent age dynamics. We find that waning immunity is the main factor explaining the return of smallpox. The fit of the mean age time series is best for this model with a high vaccine coverage, as shown by the coefficients of determination plotted in 4(d) and (e). The failed vaccine scenarios need a very long vaccine immunity duration, and a leaky vaccine model needs a very low risk of leakage to explain the data as well as the waning vaccine model with high coverage. This is demonstrated by the optimum values of the goodness-of-fit indicator *R*^2^ being closer to 1 for the waning vaccine model with high coverage, as shown in fig. 4(e).

The “lost generation” is also important in understanding the observed age structure immediately after the end of the honeymoon period. Our model shows that such a phenomenon is likely to arise if the vaccine rollout rapidly eliminates smallpox, but is also gradual enough that some children born during the honeymoon period are not vaccinated. The lost generation is however unlikely to be a key explanation for how the outbreaks could happen in the first place since, as fig. 3(a) shows, it is expected to be transient and largely disappears from the simulated epidemic dynamics after the first few post-honeymoon outbreaks.

Furthermore, we may exclude the possibility that a large number of failed, non-immunizing vaccinations led to a buildup of susceptibles. While individual cases of failed vaccinations can be documented, it is unlikely to have played a large role in epidemic dynamics, as this would have meant that a significant number of small children registered as vaccinated would get smallpox in every epidemic. This was not the case in reality, as shown in fig. 2(b) and in supplementary fig. S8. The simulations of low-coverage scenarios likewise fit the mean age time series quite poorly. A leaky vaccine might explain the mean age data reasonably well if the vaccine effectiveness is high, but as is also shown in the supplement (fig. S5), it mostly produces significantly worse fits of the age distributions of vaccinated cases. In supplementary figs. S9 and S10, we see that widespread failed vaccinations or a leaky vaccine would have resulted in smallpox becoming widespread among children again, which we do not observe.

Our simulation results show that the smallpox vaccine rollout explains the observed honeymoon period, and that it provided good protection of the vaccinated throughout childhood. The vaccine thus effectively reduced smallpox mortality in two ways: at the individual level through the vaccine immunity itself, and at the community level by securing a temporary herd immunity that caused a smallpox-free honeymoon period protecting vaccinated and unvaccinated alike.

## Discussion

We found that our model corroborates the interpretation that the vaccine rollout explains the disapppear-ance of smallpox from Copenhagen for 14 years. The model fits well the change in mean age of cases over time. It also reproduces the observed fact that the rapid disappearance of smallpox caused the rise of an immunologically näıve “lost generation” (fig. 3(a,f)) which was too old to be vaccinated in childhood, yet had never experienced the natural disease. Fits between model results and data further indicate that waning vaccine immunity was the main reason for the return of smallpox and its transformation into a disease of adulthood. A comparison with models of other possibilities, including a leaky vaccine or lower vaccine coverage, suggests that these hypotheses were likely not the cause of the smallpox resurgence. These possibilities are inconsistent with data showing that few vaccinated children were infected (figs. 2(b) and S8), and when modeled produce a quite poor fit of the mean age data.

However, the results do not reproduce the real-world dynamics exactly. As noted above, the main discrepancies in the model simulations relative to data are (1) an overestimation of cases by the model, (2) its overlooking two smaller outbreaks between 1824 and 1835 seen in data [20], (3) a slightly too high mean age of the infected before vaccination, and (4) the poor fit between the model’s predicted age distributions of unvaccinated infections and observations. As for (1), any overestimation of the number of infections is likely to be due to two factors: for one, it is a known problem with SEIR-type models that they overestimate the number of infections [39]. Furthermore, our model ignores efforts to suppress the outbreaks through non-pharmaceutical interventions such as quarantine. We know that such measures were taken in the period between 1824 and 1835.

We also did not consider possible seasonality. This may otherwise have suppressed the epidemics at certain times of year, possibly explaining deviations from the predicted semi-regular 6-year cycle of our model. We believe this, possibly in combination with our neglecting mitigation efforts, to be the main reason why the model oversimplifies the complex outbreak dynamics of the 1820’s and 30’s (discrepancy (2)).

Regarding the high pre-vaccination mean age at infection (discrepancy (3)), literature provides different possible estimates for the true value. Duncan et al. [40] estimate this age to be between two and five years, while Davenport et al. [41] gives a mean age at death of 7.8 years for London (St Martin-in-the-Fields) in the mid-1700s, decreasing to 3.9 years by the end of the century. This puts the mean age given by the simulation without vaccination (about 7 years) in the high end of the realistic range. A likely reason for this overestimation is the fact that we neglect social structure and contact heterogeneities in our model. If we had included such heterogeneities, such that for example children interacted more with others of the same age, this might have led to smallpox spreading even more readily among the youngest age groups given their high susceptibility. This would lead to a lower mean age of infected in the endemic pre-vaccination state. A lower starting point for the mean age of infected would similarly improve the mean age fit, as the present version of the model tends to overshoot the earliest mean age data points.

The poor fit between the unvaccinated age and birth-year distributions of cases predicted by the model and those observed in data (discrepancy (4)) may in part be due to our assumptions about age at vaccination and the data on which we base them (fig. S2 and [16]). These data are from Frederiksborg County, located close to but outside of Copenhagen, and so may not reflect the situation in Copenhagen itself. The data show a quickly declining age at vaccination and indicate that 80 % of children were already vaccinated by their first birthday in 1825. This, we also use to parameterize our model. If this observation does not match the situation in Copenhagen, and there were more unvaccinated young children present, this may explain the discrepancy of the age and birth year distributions. In addition, the data are not granular enough to show whether the 0-1-year olds were vaccinated early or late in their first year of life, which may have significantly affected the number of cases in the youngest age group.

As a final caveat should be noted that our model takes population size to be constant for the sake of simplicity, although from censuses we see that the population almost doubled during the period [42]. Whether this would change the behaviour of the disease depends on one’s assumptions about the dependence of infectivity on population size and density. In our model we assume that infectivity is dependent on population density, and we correspondingly divide the infection terms *βSI* by the population size *N*. Therefore, we would not expect the model’s disease dynamics to change greatly if we were to incorporate population growth, unless coupled with a simultaneous population density growth. The effect of population growth on density, and thus on disease dynamics, is further complicated by the historical situation in Copenhagen at the time. As the city greatly expanded geographically during the period after the abandonment of its medieval defenses [43], the effect of its population growth on density is not straightforward.

Despite the caveats described here, we believe that lessons may be drawn from our model, since the crucial dynamics of varying immunity as a function of age and the different sizes of age groups are taken into account, and since the model fits the changing mean age of the smallpox cases and their birth-year distributions well.

It is worth commenting on the quality of available data. Between May 1835 and the 1860’s, data on infections are affected by underreporting of mild cases or missing entirely, since smallpox was no longer considered a serious public health risk and not subject to mandatory quarantine. This change particularly affects the data which we studied for the 1835 outbreak, as the hospitalization policy was changed in May that year [13]. Thus, the data only cover the period from May to December, only relatively severe cases, and only those hospitalized in one particular hospital. Reporting on the health condition of the population has been required of Danish physicians since 1803 [44], but it is only from 1867 onward that we have reasonably complete and digitized data [21]. It is therefore possible that the graphs of figs. 2(a) and 4(b) miss significant epidemic events in the intervening years, as they show only mortality data. This is especially true if the CFR of the outbreaks was low, as is likely in the vaccine era [45].

An approach to verifying our estimates of the durability of the immunity conferred by the original cowpox-based vaccine would be to look at later studies of vaccine effectiveness. Serological studies tend to indicate that smallpox vaccination provides lifelong antibody protection against the disease, while T-cell responses decline with a half-life of 8-15 years [46]. The US CDC fact sheet on the vaccinia-based smallpox vaccine states that waning of transmission blocking immunity is expected after 3-5 years [37]. However, this may represent a cautious estimate, as it is intended as a guideline for the public regarding when boosters should be considered. According to Arita [35], Dixon [34] estimated an 87.5 % vaccine effectiveness after 10 years and 50 % after 20 years. These numbers are mentioned in the context of effectiveness against infection, but this is not stated outright. Finally, another historical study by Nishiura & Eichner [36] estimates an exponentially declining vaccine effectiveness with a half-life of roughly 60 years against death and 35 years against severe infection. They provide no results for mild cases. Observations from 19*^th^* century Copenhagen likewise indicate that the protection conferred by the early cowpox-based vaccine against infection was not lifelong. Based on our results we estimate a duration of immunity of 20-30 years, including against mild infections. This is consistent with the Dixon and Nishiura & Eichner studies.

One important question to ask is whether the smallpox dynamics in Copenhagen after vaccination as explored here can be generalized to other contexts. Have similar dynamics recurred in other locations and times with a similar vaccination background? And, perhaps most importantly, do we have data from other places in a comparable situation which reproduce the dynamics? We believe that Scotland in the mid-19*^th^* century might provide such a comparable example. Scotland rapidly rolled out the vaccine after its invention, but towards 1850 the uptake had decreased to as little as 20 %. Therefore, it was decided to make vaccination mandatory in 1863 [47]. This led to a sharp increase in vaccination coverage to over 80 % in later birth cohorts. Correspondingly, we should expect to see a shift in the ages of smallpox victims in Scotland as children are immunized and the disease increasingly affects non-immunized adults or those with an old vaccination. Judging from data from a report to the British Parliament by the late-1800’s Vaccination Commission, this is indeed what happened [48]. Before 1863, most people who died from smallpox were less than five years old. In the 20-year period after vaccination became mandatory, most smallpox victims were in their 20’s and 30’s, while overall smallpox mortality was greatly reduced. Based on available data we cannot know if there was a generation of people who were neither vaccinated nor naturally immune in Scotland as was the case in Denmark. However, the shift of smallpox from a childhood disease to a disease of adulthood happened with about the same timing relative to the vaccination rollout in Scotland as it did in Copenhagen half a century previously. Given this, we believe that we can conclude that it was indeed the vaccination rollout which caused the age shift described here, and not other historical factors present around 1800.

In conclusion, the vaccine against smallpox greatly reduced the mortality due to the disease wherever it was adopted. We have used data from Copenhagen to get a glimpse into the resulting changes in disease dynamics, and have thereby uncovered some details of how the very first vaccine worked. To summarize our results, we find that the model which best explains the smallpox dynamics observed in Copenhagen after the introduction of the vaccine is one including a waning vaccine immunity with a mean duration of 20-30 years. On the other hand, our assembled data and modeling results contradict the hypotheses that smallpox resurgence was caused by failed vaccinations or vaccine leakage. In addition, we see in the Copenhagen case data that the generation born during this honeymoon period, particularly before mandatory vaccination, were highly vulnerable to later infection and are overrepresented in the case statistics of the first epidemics after the return of smallpox. Our findings thus highlight one way in which a childhood vaccination campaign can cause the demographics of a disease to shift radically. In recent history, this has also been predicted and observed for Rotavirus disease following its vaccine introduction [49, 50], and something similar is currently happening with pertussis [51]. All this is an example of how historical data may still be relevant to present-day epidemiology. It shows how they may be combined with modern methods to produce new results and to better understand the phenomenon of honeymoon periods and epidemic recurrence under current vaccination programs. In the present day, we are experiencing both the end of a pandemic and the resurgence of some childhood infectious diseases in the developed world [52]. In this context the example of smallpox in the era of early vaccination efforts - and nearly 200 years before its eradication globally-provides an invaluable opportunity to study how a disease behaves for decades after the introduction of a vaccine.

## Author contribution statement

LS conceptualized the study, acquired funding, administered, and supervised the project. SKP located and curated the data. AE created the software for the numerical model, performed the analysis and visualized the results. LS, KS, and AE devised the theoretical methodology in collaboration, while BG and KS provided input on modeling and result presentation, and SKP provided historical insights and methodological advice. LS and AE wrote the manuscript, and all authors reviewed and approved it.

## Data availability

The code used to simulate the smallpox dynamics and generate the plots shown here and in the supplement is available on Figshare (https://doi.org/10.6084/m9.figshare.28182581). Data on vaccination coverage, case counts, and vaccination statuses are assembled from the medical journal Bibliotek for Læger, the Danish National Archive (Rigsarkivet) [19, 20, 53], and contemporary publications [11–13, 22]. The aggregated hospitalization data can be found on GitHub (https://github.com/basspoder/smallpox_post_ honeymoon_phase_1824_1872/releases/tag/Dataset).

## Supporting information

Supplementary sensitivity and goodness-of-fit analyses

## Acknowledgements

This project received funding from the Danish National Research Foundation (DNRF170) and NordForsk (project no. 104910). We thank the other members of the PandemiX Center at Roskilde University, the USYS at the ETH Zürich, and the EEB at Princeton University, for many enlightening discussions.

## AI use statement

The Claude Sonnet 4.6 LLM was used to provide an estimate of death and immigration rates to reproduce the observed demography as described in the Methods section.

## Notes

### Competing Interest Statement

The authors have declared no competing interest.

### Author Declarations

The Danish National Archives (Rigsarkivet)

### Summary of Updates

A major revision of the main manuscript and supplement following review. Now hypotheses explaining the return of smallpox other than vaccine immunity waning are thoroughly tested, and we evaluate the goodness-of-fit of the model using the coefficient of determination. Finally, additional data for a smallpox outbreak in 1828-30 are now studied in more detail.

