## Supplementary sensitivity and goodness-of-fit analyses for "Modeling epidemiological patterns of smallpox in Copenhagen in the 19*^th^* century after the introduction of the vaccine"

### Supplementary material for the article "Modeling epidemiological patterns of smallpox in Copenhagen in the $19^{th}$ century after the introduction of the vaccine"

June 15, 2026

This supplement provides additional details on the reasoning behind our assumptions about the age at vaccination, demography of Copenhagen in the $19^{th}$ century and in our model, the parameters relevant for simulating demographic processes, and our assumptions about the vaccine waning time distribution. In addition, we provide further quantification of the goodness-of-fit between our model given various assumptions and the age distribution data and observed honeymoon period duration. To allow the reader to inspect the effect of changing these assumptions on outbreak frequency and age distribution, we include plots of the number of infections and age patterns over time. As our choice of $R_0$ is slightly uncertain, we test the sensitivity of our results to variations in this parameter. Finally, we provide additional information on the smallpox outbreak of 1871-72, which is not covered in detail in the main manuscript, but which does show up in time series and mean age data.

#### Extended model diagram

In the interest of legibility, we have cut some details on model assumptions about age at vaccination from the model diagram in the main manuscript. Here, we include the full diagram (fig. S1) including the details on age at vaccination as a function of time.

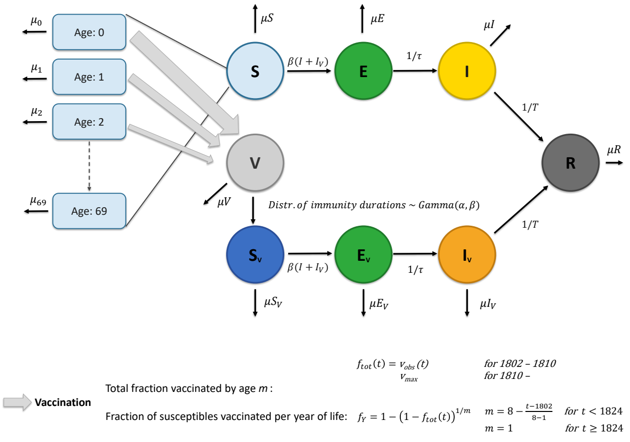


Figure S1: An extended version of the diagram presented in the main manuscript, including details on the assumptions about the age at vaccination over time. Births and immigration are not shown.

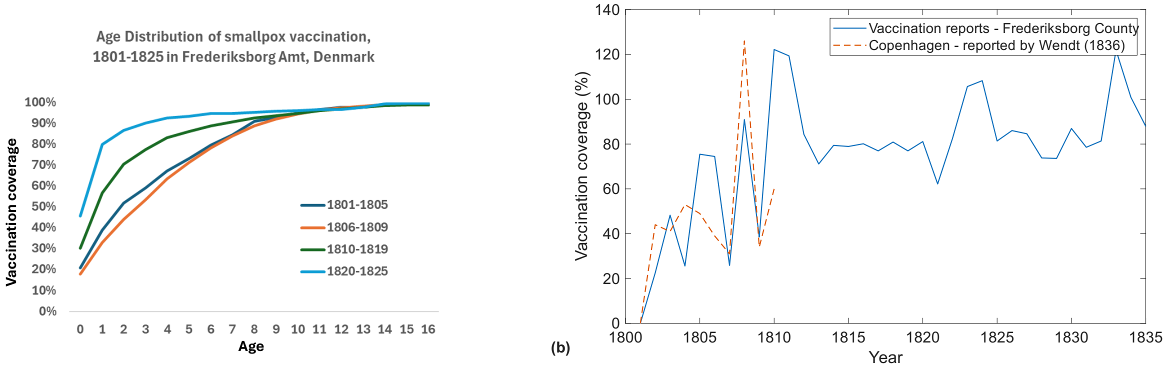


Figure S2: (a) A plot showing cumulative vaccination coverages as a function of age for selected periods during the early 1800's. (b) A plot of total vaccination coverage in Frederiksborg County, calculated by dividing the number of vaccinations by the birth cohort size of the previous year, adjusted for a 10 % mortality in early infancy. Dashed line shows data from Copenhagen, reported by Wendt [9].

### Age at vaccination

The assumptions made about the age at which children were vaccinated are based on data from [7, 9]. In fig. S2(a), we display a figure showing the cumulative vaccination coverage as a function of age at different time points between 1801 and 1825. The cumulative coverages are calculated in the same way as the coverage time series displayed in the main manuscript, i.e., by dividing the number of people vaccinated at a given age with the size of the corresponding birth cohort, corrected for a 10 % rate of infant mortality before vaccination. Panel (b) shows a time series of the total vaccination coverage in Frederiksborg County between 1801 and 1835, and for Copenhagen from 1801 to 1811, adjusted for a 10 % mortality in early infancy.

### Age-dependent mortality and migration rates

The mortality rates $\mu_i$ are estimated by using a realistic infant mortality based on data from the Human Mortality Database [1], increased from 0.15 to 0.18 to account for the urban penalty (the value given by the reference is for all of Denmark, i.e., also rural areas). The remaining demographic parameters were "guessed" by letting the Claude (Sonnet 4.6) LLM estimate a set of parameters leading to a demography resembling the real one provided by [8]. We subsequently verified that these parameters resulted in the correct population age structure and manually adjusted them as needed. A plot of the resulting age structure versus the observed one can be seen in fig. S3.

The age-dependent mortality rates used were as follows:

$$
\begin{aligned}
&\mu_i = [0.1800, 0.050, 0.050, 0.050, 0.0130, 0.0080, 0.0070, 0.0060, 0.0060, 0.0060, \ldots \\
&0.0050, 0.0050, 0.0050, 0.0060, 0.0060, 0.0070, 0.0080, 0.0090, 0.0100, 0.0220, \ldots \\
&0.0240, 0.0240, 0.0260, 0.0260, 0.0280, 0.0280, 0.0294, 0.0308, 0.0322, 0.0338, \ldots \\
&0.0177, 0.0185, 0.0194, 0.0203, 0.0213, 0.0223, 0.0234, 0.0245, 0.0256, 0.0269, \ldots \\
&0.0281, 0.0295, 0.0309, 0.0323, 0.0339, 0.0355, 0.0372, 0.0390, 0.0408, 0.0428, \ldots \\
&0.0448, 0.0469, 0.0492, 0.0515, 0.0540, 0.0565, 0.0592, 0.0620, 0.0650, 0.0681, \ldots \\
&0.0713, 0.0747, 0.0783, 0.0820, 0.0859, 0.0900, 0.0943, 0.0988, 0.1035, 0.1035]/year \quad (1)
\end{aligned}
$$

The index $i$ here indicates the age group in question, from 0 to 69. Migration, $\Lambda_i$, is neglected for all age groups except for the 15-25-year-olds, since their demographics are clearly strongly affected by migration. We set the migration rates to be $\Lambda_{15-25} = 0.35$ individuals/day. Unfortunately, we do not have data available to back up this estimate, but must resort to choosing a value that best fits the demography.

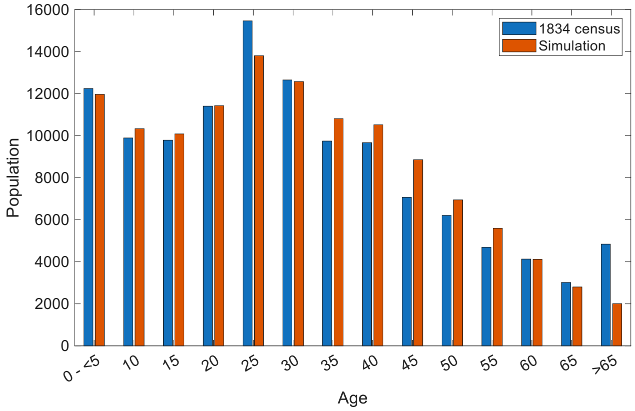


Figure S3: Bar graph of the age structure of the population of Copenhagen in 1834 (blue bars) compared to that used in our model (orange bars). Numbers on the x-axis show the upper end of the agebin in question, with all +65-year-olds included in the same agebin.

### Preliminary simulation

To obtain the distribution of immunity prior to vaccination, we run the simulation for 70 years with no vaccination. We then load the resulting immune populations in each age group, $R_i$, into the post-vaccine simulation and use it as a starting condition. The timeseries of the pre- and post-vaccination epidemic dynamics can be seen in fig. S4. Here, it is compared with the observed mortality from smallpox between 1750 and 1880. The model is not able to capture the apparent erratic smallpox outbreaks in 1770-1800. In a previous work, Eilersen discusses possible causes of these dynamics, which are in general difficult to capture with compartmental disease models [3].

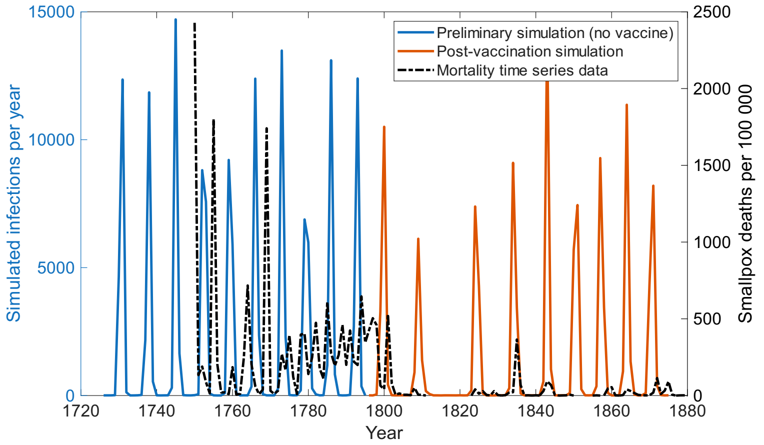


Figure S4: Timeseries of the simulation before vaccination rollout (blue) and after rollout (orange).

### Fitting models with leaky vaccine and limited coverage

In fig. S5 we show plots of the coefficient of determination of the fit of our model to age distribution data for various assumptions including different coverages and whether the vaccine is waning or leaky. It should be noted that age distribution data are different from birth-year distribution data, as the age distributions are not only affected by immunity but also by the timing of the outbreaks and interval between them. In general, the goodness-of-fit for any of the models is much better for the vaccinated age distributions than for the unvaccinated ones ($R^2$ closer to 1). A high vaccination coverage also gives the best fits for the age

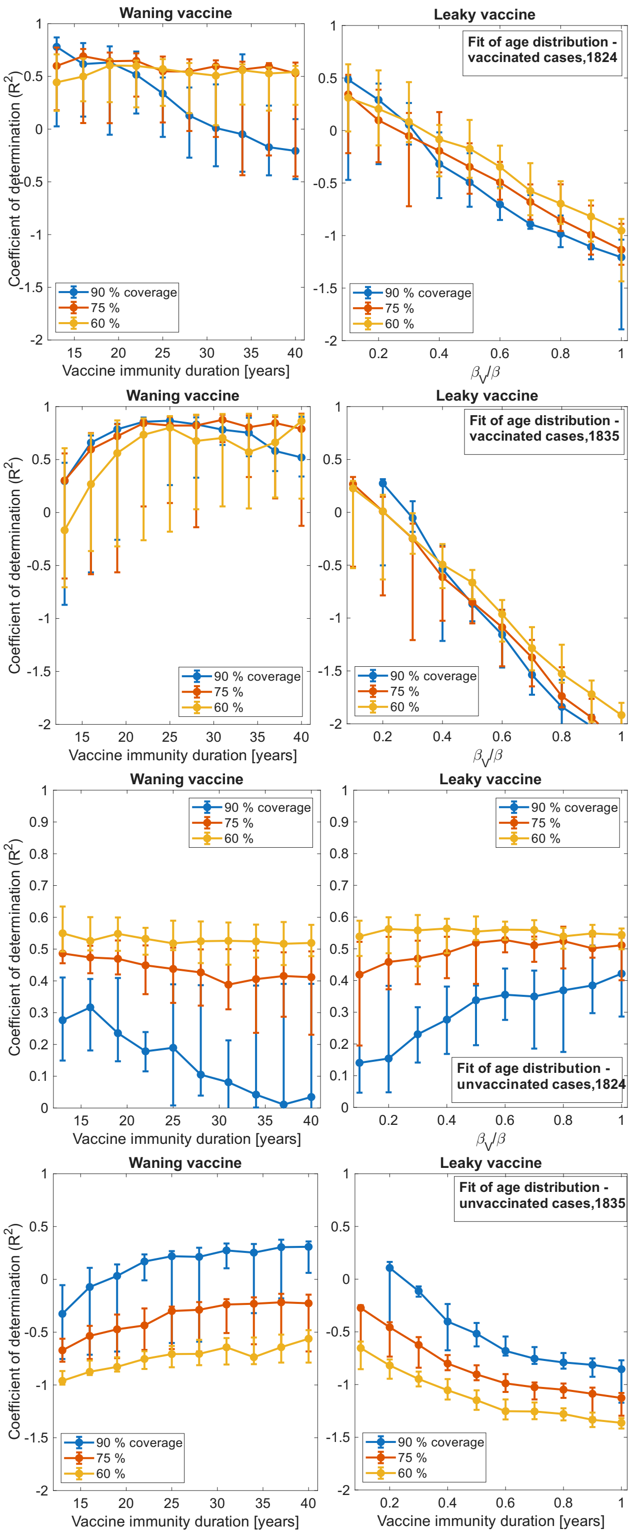


Figure S5: Plots of the goodness-of-fit as measured by the coefficient of determination $R^2$ for the simulated age distributions against the observed age distributions of vaccinated and unvaccinated cases in 1824 and 1835. From top: vaccinated cases, 1824; vaccinated cases, 1835; unvaccinated cases, 1824; unvaccinated cases, 1835. The left column shows $R^2$ values for the waning vaccine model, while the left column shows thos for a leaky vaccine model. Errorbars show 95 % confidence intervals. In the leaky vaccine model, we vary the relative transmission rate $\beta_V/\beta$ of the vaccinated, while for the waning vaccine model we vary vaccine immunity duration. Coverages are varied for both models.

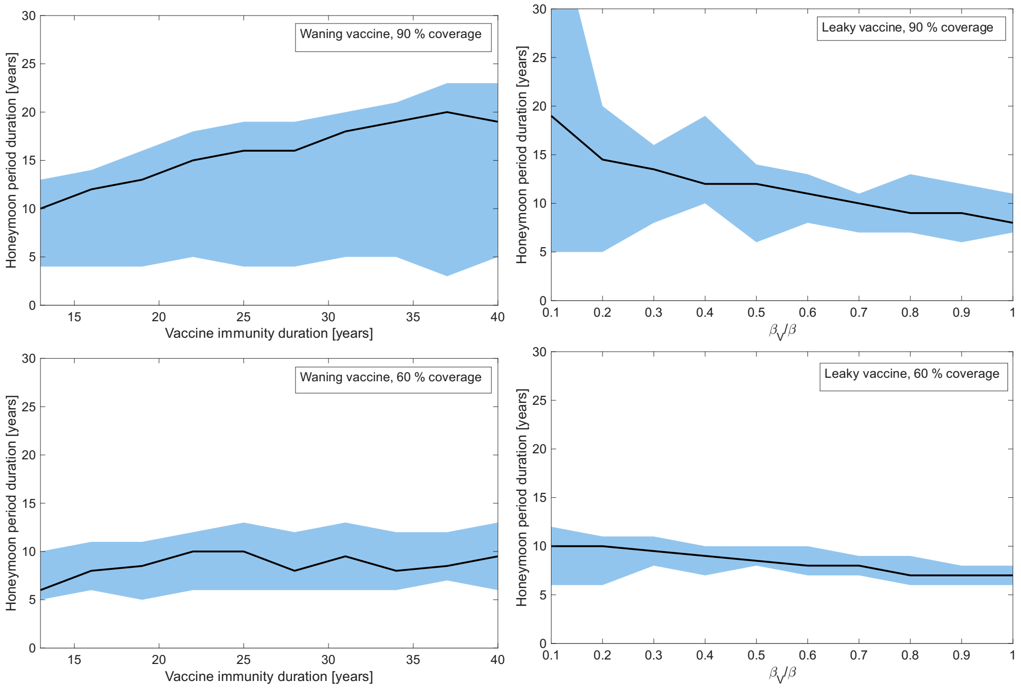


Figure S6: Honeymoon period durations as predicted by the model for varying maximum vaccine coverage $v_{max}$, vaccine immunity duration, and reduction of $\beta$ in the vaccinated given a leaky vaccine. Black lines show the median, shaded blue area show the 95 % confidence intervals. Compare the observed honeymoon period duration of 14 years.

distribution of unvaccinated cases in 1835, and the worst for the unvaccinated and partly for the vaccinated age distribution in 1824. This may be due to the high-coverage models overestimating the interval between epidemics, which also changes the age distributions. The correct honeymoon period duration is only recreated by the highest vaccine coverage. In most cases, the waning vaccine model does better than the corresponding leaky vaccine model, depending of course on the exact choice of values of the variable parameter (vaccine immunity duration for the waning vaccine model and infectivity relative to the unvaccinated, $\beta_V/\beta$, for the leaky vaccine model).

In fig. S6 we similarly show the honeymoon period duration predicted given the same modelling assumptions. We see that the model only is able to reproduce the observed honeymoon period duration of 14 years for a high vaccine coverage of 90 %. In the leaky vaccine model, a reduction in infectivity of $\beta_V/\beta = 0.2$ is necessary to give the correct duration, while for the waning vaccine model, an immunity duration of 20 years gives the most accurate duration. We believe these results to be a good argument against the prevalence of failed, non-immunizing vaccinations. We also conclude based on fig. S5 that vaccine immunity waning is the most likely explanation for the observed age distributions, albeit with slightly less certainty.

In fig. S7, we show a comparison of the simulated *birth year distributions* of the unvaccinated cases in 1828-30 and 1835, which were left out of the main manuscript. Note that these are birth year distributions, and thus different from the *age distributions* used to calculate the $R^2$ values shown above. The big spike in panel S7(a) is an artifact of the model. It is due to the fact that we reintroduce smallpox into the migrating young-adult age groups. As there are very few cases in the simulation between 1828 and 1830, these reintroductions skew the age distribution.

The birth year distributions can be compared and contrasted with fig. S8, which shows the observed *age distributions* of cases in the vaccinated and unvaccinated. Particularly notable is the observation that very few vaccinated children were infected, indicating that there were few children with ineffective ("failed") vaccines and the vaccine did not leak. Neither was there any noticeable waning in the first years after vaccination. In

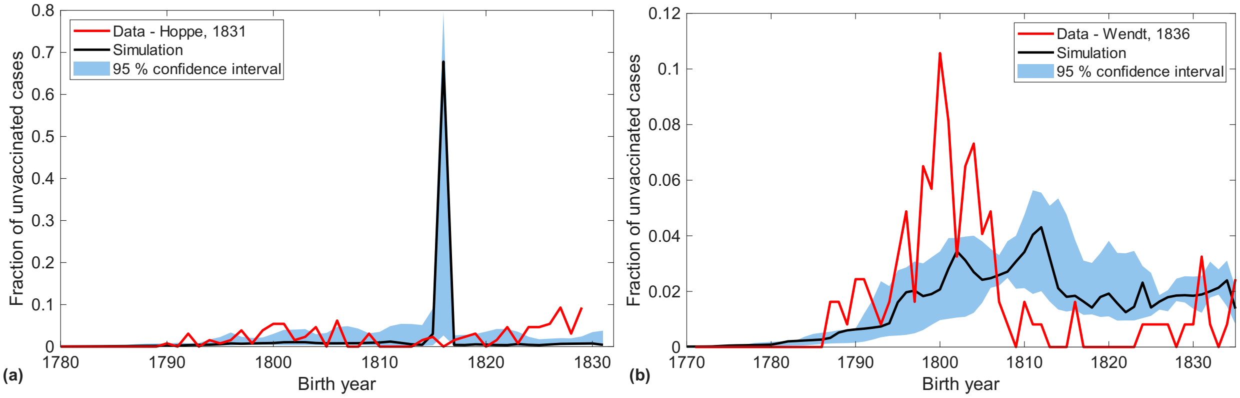


Figure S7: Birth year distributions of unvaccinated cases compared with simulation predictions. (a) shows the distribution from the 1828-30 period (red line) and the simulated distribution (black line). (b) shows the same, but for 1835.

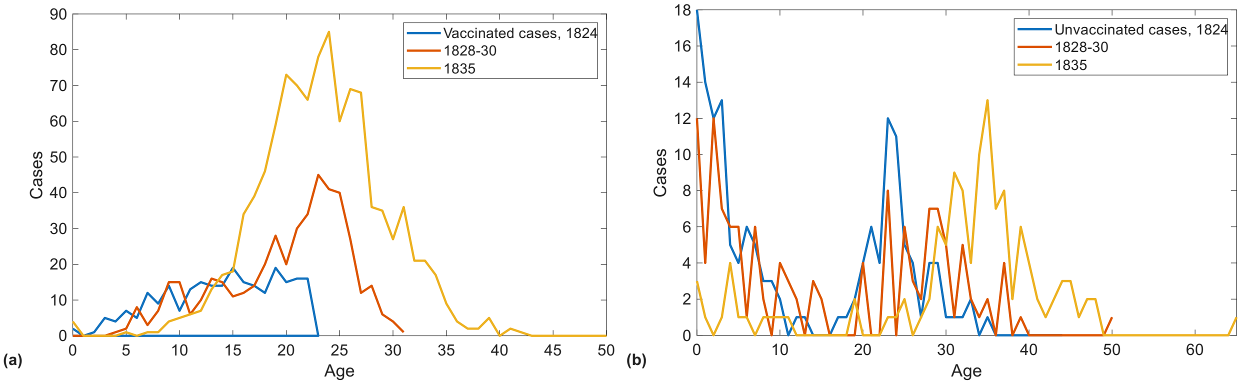


Figure S8: *Age distributions* of (a) vaccinated and (b) unvaccinated cases, as opposed to birth year distributions. Note the very few cases in vaccinated children, and the decreasing number of cases in unvaccinated children in later outbreaks.

the unvaccinated (panel (b)), one observes that in 1824 and to a lesser extent 1828-30, cases in the youngest unvaccinated children were common, but this was strongly reduced by 1835. This indicates that the tendency to vaccinate children earlier and earlier as observed in fig. S2 effectively prevented smallpox in the youngest age groups.

### Age distributions using alternative model hypotheses

To illustrate the difference in age patterns generated by assuming a leaky vaccine or many failed vaccinations, we here show age distributions of infections in the vaccinated and unvaccinated over time given a low vaccine coverage of 60 % (fig. S9) or a leaky vaccine that only protects 80 % against infection, but does so permanently (fig. S10). We see that in both cases, smallpox becomes more prevalent in the youngest age groups than for the waning vaccine of the main manuscript, which contradicts observations. The model assuming a leaky vaccine with good protection against infection has fewer cases in children. On the other hand it predicts a frequency of outbreaks that is much lower than in reality, with only four post-honeymoon outbreaks.

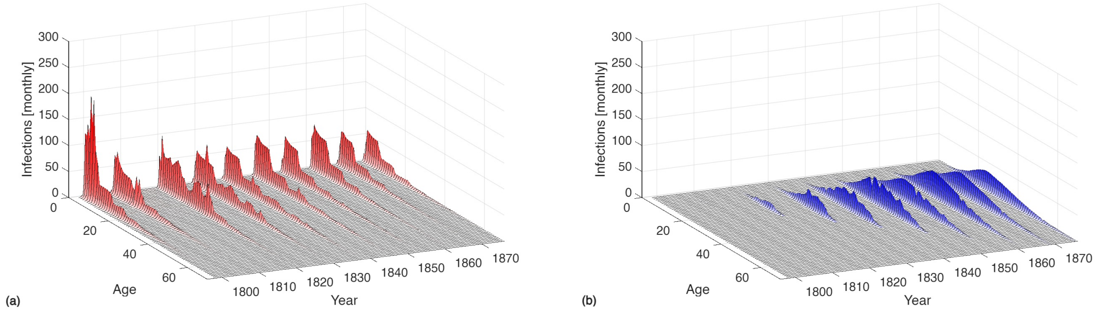


Figure S9: Age distribution of monthly infections over time in (a) vaccinated and (b) unvaccinated people, assuming a low vaccination coverage of 60 % (or alternatively many failed vaccinations).

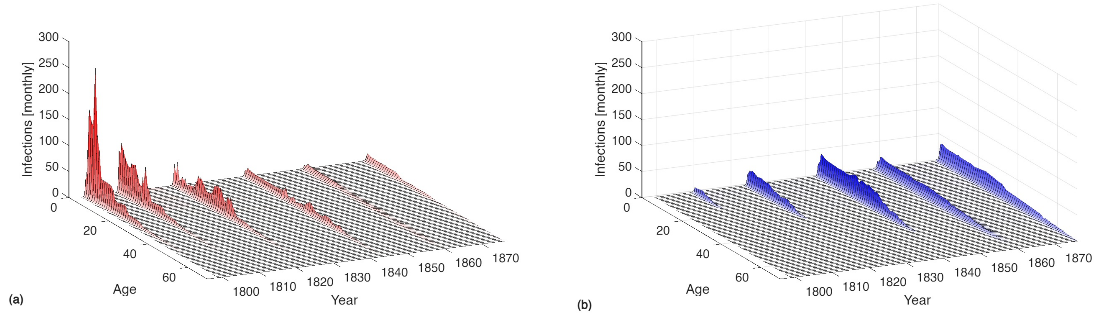


Figure S10: Same as fig. S9, but for a leaky vaccine giving a protection of 80 % against infection and a coverage of 90 %.

### Sensitivity of the model to variations in $R_0$

In fig. S11, we show the effect of varying $R_0$ on outbreak frequency and mean age of the infected. Changing $R_0$ from 5 to 7 yields more frequent epidemics and a lower mean age of the infected in general. The magnitude of these effects is only modest, and we therefore do not believe that a small change in our assumptions about the $R_0$ of smallpox at the time would fundamentally affect our conclusions.

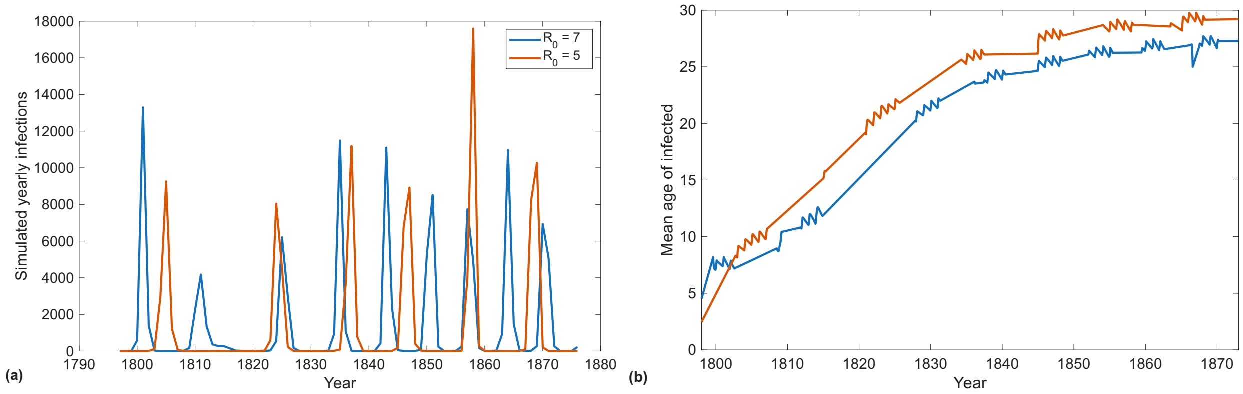


Figure S11: (a) Example time series when varying $R_0$. (b) The evolution in mean age of the infected over time when varying $R_0$

### Assumptions about the distribution of vaccine waning times

The assumption that vaccine waning times are Gamma distributed is a deviation from the assumption of exponentially distributed waiting times that is very common in SEIR models. We believe it to be justified, as it is in good agreement with the data in fig. 2(d) of the main manuscript, which shows that very few cases occur in the most recently vaccinated when compared to populations vaccinated further in the past. Additionally, it fits with observational data for some viral diseases [5] and with the assumption of a protective threshold for antibodies [4]. Finally, for smallpox we observe a limited variability of incubation and infectious period duration [2]. As the latent, incubation, and infectious periods of smallpox have a well-defined and consistent duration, it seems likely that the duration of immunity against the disease would likewise be consistent.

### Information on the 1871-72 outbreak

Fig. S12 shows observed hospitalizations and deaths per 1000 inhabitants in Copenhagen during the 1871-72 smallpox outbreak, which was part of the 1870-75 pandemic [6]. The hospitalization policy was vastly different in this outbreak in comparison with those before 1835; only the most severe cases were hospitalized in 1871-72, whereas in 1824-35 all infected were hospitalized for containment reasons [9]. Due to this large difference, it makes little sense to directly compare this outbreak to those in the main article, and this plot is therefore only included in the supplement. Nonetheless, it is instructive to compare the plot in fig. S12, which shows that smallpox hits the 40-60-year-olds hardest, to those of the 1824-35 outbreaks where young adults are hit hardest, and finally to contrast both of these to the pre-vaccination status of smallpox as a childhood disease.

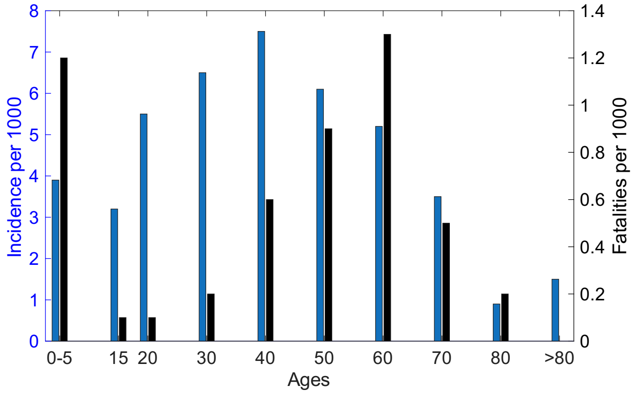


Figure S12: Rates of hospitalizations (blue, left y-axis) and fatalities (black, right y-axis) per thousand inhabitants in Copenhagen in the outbreak 1871-72. Note that the criteria for hospitalization were much stricter in this outbreak than before 1835, meaning that the hospitalization rate cannot be taken to accurately reflect disease prevalence.


[5] C. Hernandez-Suarez and E. Murillo-Zamora. Waning immunity to sars-cov-2 following vaccination or infection. *Frontiers in Medicine*, 9:972083, 2022.

[6] J. D. Rolleston. The smallpox pandemic of 1870-1874:(section of epidemiology and state medicine). *Proceedings of the Royal Society of Medicine*, 27(2):177, 1933.

[7] Several. Vaccinationsprotokol [vaccination protocol]. Rigsarkivet, 1800-1914.

[8] Several. Folketælling 1834, københavn [1834 census, copenhagen]. Rigsarkivet, 1834.

[9] J. C. W. Wendt. Bidrag til børnekoppernes og vaccinationens historie i danmark, og om de sidste herskende koppe-epidemier. *Unknown*, 1836.
